# Particulate matter air pollution and COVID-19 infection, severity, and mortality: A systematic review

**DOI:** 10.1101/2022.11.16.22282100

**Authors:** Nicola Sheppard, Matthew Carroll, Caroline Gao, Tyler Lane

## Abstract

**Background and objective:** Ecological studies indicate ambient particulate matter ≤2.5mm (PM_2.5_) air pollution is associated with poorer COVID-19 outcomes. However, these studies cannot account for individual heterogeneity and often have imprecise estimates of PM_2.5_ exposure. We review evidence from studies using individual-level data to determine whether PM_2.5_ increases risk of COVID-19 infection, severe disease, and death.

**Methods:** Systematic review of case-control and cohort studies, searching Medline, Embase, and WHO COVID-19 up to 30 June 2022. Study quality was evaluated using the Newcastle-Ottawa Scale. Results were pooled with a random effects meta-analysis, with Egger’s regression, funnel plots, and leave-one-out and trim-and-fill analyses to adjust for publication bias.

**Results:** *N*=18 studies met inclusion criteria. A 10μg/m3 increase in PM_2.5_ exposure was associated with 66% (95% CI: 1.31-2.11) greater odds of COVID-19 infection (N=7) and 127% (95% CI: 1.41-3.66) increase in severe illness (hospitalisation or worse) (N=6). Pooled mortality results (N=5) were positive but non-significant (OR 1.40; 0.94 to 2.10). Most studies were rated “good” quality (14/18 studies), though there were numerous methodological issues; few used individual-level data to adjust for confounders like socioeconomic status (4/18 studies), instead using area-based indicators (12/18 studies) or not adjusting for it (3/18 studies). Most severity (9/10 studies) and mortality studies (5/6 studies) were based on people already diagnosed COVID-19, potentially introducing collider bias.

**Conclusion:** There is strong evidence that ambient PM_2.5_ increases the risk of COVID-19 infection, and weaker evidence of increases in severe disease and mortality.

**Funding:** This review was completed as a Scholarly Intensive Placement project by NS, which received no funding.

**Competing interests:** The authors declare no competing interests.

**Registration:** This study was registered on PROSPERO on 8 July 2022 (CRD42022345129): https://www.crd.york.ac.uk/prospero/display_record.php?ID=CRD42022345129

## Introduction

A considerable body of ecological evidence suggests fine particulate matter air pollution ≤2.5μm (PM_2.5_) increases the risk of COVID-19 infection, severity, and death (1–3). However, this evidence relies on comparisons of geographic units, which do not account for individual-level differences and often misclassify exposures due to poor precision/resolution in PM_2.5_ estimates (3). Associations between PM_2.5_ and COVID-19 may therefore be spurious, confounded by socioeconomic differences that influence exposure to air pollution and COVID-19 risks (4).

Nevertheless, are several reasons to suspect that PM_2.5_ increases COVID-19 risks. PM_2.5_ increases expression of Angiotensin-Converting Enzyme 2 (ACE2), which the COVID-19 spike protein uses to bind to and enters host cells (3,5). Though there is limited evidence for ambient PM_2.5_, studies of cigarette smoking suggest it inhibits cell defence against infections (5). PM_2.5_ and COVID-19 may also operate in tandem, both independently worsening respiratory and cardiovascular health, leading the combination of exposures to increase the likelihood of severe disease and death (3,6).

This systematic review builds on previous reviews (1–3) by focusing on studies using individual-level data that can provide more precise exposure estimates and better account for confounders. We address the following questions:

1. Is ambient PM_2.5_ exposure predictive of COVID-19 infection, severe COVID-19 disease, and COVID-19 mortality?
2. Do discrete high PM_2.5_ events like wildfires predict COVID-19 infection, severe COVID-19 disease, and COVID-19 mortality?

## Methods

This review is registered on PROSPERO (7) and is reported according to PRISMA 2020 guidelines (8). A completed PRISMA checklist is available on a public repository (9).

### Inclusion/exclusion criteria

To be eligible for inclusion, studies had to analyse individual-level data on the association between PM_2.5_ and COVID-19 infection, severity, or mortality using either a case-control or cohort design. Studies needed to present original research in an English-language peer-reviewed journal no later than 30 June 2022.

Studies were ineligible if they used ecological, cross-sectional, case-series, animal, or in-vitro designs; studies with a mixture of methods that included either case-control or cohort design were considered eligible. Hypothesis, review, editorial, commentary, and opinion pieces were excluded, as were pre-prints and conference presentations. Studies not using PM_2.5_ or only examining indoor air pollution or tobacco smoke as the pollutant exposure were excluded.

### Search strategy and screening

We searched Medline, Embase and the World Health Organization COVID-19 database using terms listed in the Appendix. In addition, we screened the reference lists of grey literature and previous systematic reviews on similar topics for studies meeting the inclusion criteria. Two study authors (NS & TL) independently screened abstracts and full-texts for eligibility. Disagreements were resolved between screening authors or, failing that, by a third author (MC).

### Data extraction and quality assessment

Two authors (NS & TL) independently extracted data and assessed study quality, and a third author (MC) settled disagreements. Data extraction focused on characteristics of the study sample/population, operationalisation of PM_2.5_ measurement, and COVID-19 outcomes. Effect size and direction, coefficient type (e.g., Hazard Ratio, Odds Ratio), and confidence intervals were tabulated.

Quality was assessed using the Newcastle-Ottawa Scale (NOS) (10) and scores were converted to Agency for Health Research and Quality (AHRQ) standards using the rubric in Shamsrizi et al (11):

- **Good quality**: 3 or 4 stars in *Selection* domain AND 1 or 2 stars in *Comparability* domain AND 2 or 3 stars in *Outcome* domain
- **Fair quality**: 2 stars in *Selection* domain AND 1 or 2 stars in *Comparability* AND 2 or 3 stars in *Outcome* domain
- **Poor quality**: 0 or 1 star in selection domain OR 0 stars in *Comparability* domain OR 0 or 1 stars in *Outcome* domain

### Meta-analysis

Results were pooled using a random-effects meta-analysis with the *metafor* (12) and *metaviz* (13) packages in R (14). A Meta-analysis Of Observational Studies in Epidemiology (MOOSE) (15) checklist is available in our public repository, along with meta-analysis code and data (9). Studies were limited to those rated “good” or “fair”, with sensitivity analyses including all studies regardless of quality. Assuming inherent variance due to differences in populations and methods, we used random effects models and report the *I*^2^ statistic for heterogeneity. All outcomes were converted to Odds Ratios for synthesis. Egger’s regression and funnel plots tested for publication bias. While not specified in the original protocol on PROSPERO, we added trim-and-fill and leave-one-out sensitivity analyses to test the robustness of results.

Where studies reported multiple outcomes, we prioritised the following: lengthiest PM_2.5_ measurement; most comprehensive measure of outcomes (e.g., serology and self-reported symptoms rather than one or the other; hospitalisation+ rather than just hospitalisation or ICU admittance); complete rather than restricted samples/populations (e.g., analysis of the entire Ontario population rather than only test-takers in Sundaram et al (16)); models adjusting for socioeconomic factors; and the indicator of “least” severity (e.g., hospitalisation over ICU admittance (17)); continuous PM2.5 measures (only one study used a non-continuous measure). For the two studies by Mendy et al., we used only the more recent, larger study (18) since it included all participants from the earlier one (19). This approach to outcome selection was not specified in the protocol as outcome reporting preferences of studies were unforeseeable.

## Results

### Search results

Search strategy results are in Figure 1 below. The initial literature search of Medline, Embase and the WHO COVID-19 database yielded 1,442 studies, which was reduced to 18 after screening. One study was excluded even though it met the inclusion criteria because it reported only statistically significant results rather than all results regardless of significance (20). A full list of screened studies along with reasons for exclusion is available in our public repository (9).

**Figure 1.**
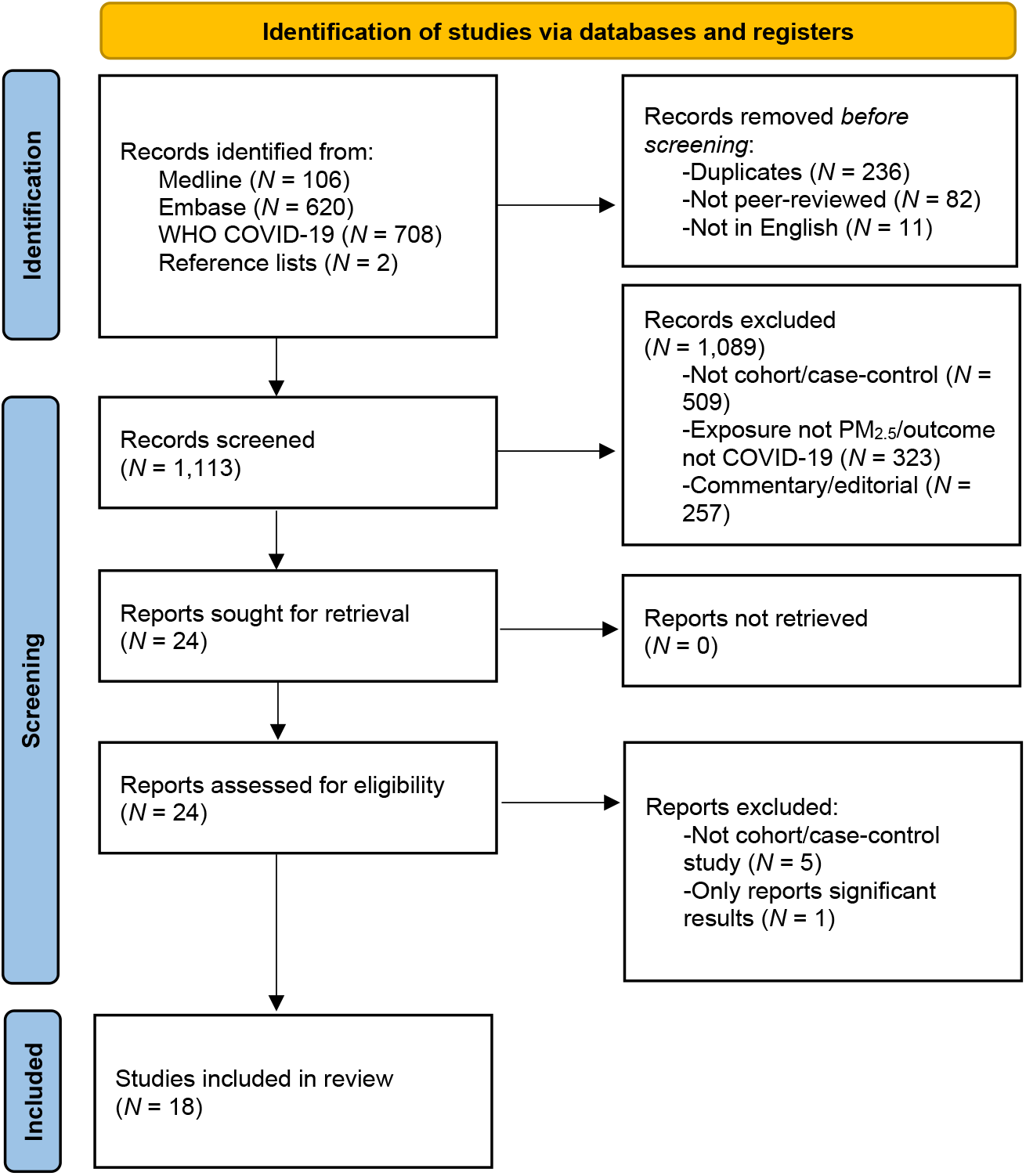
Screening flow diagram

**Figure 2.**
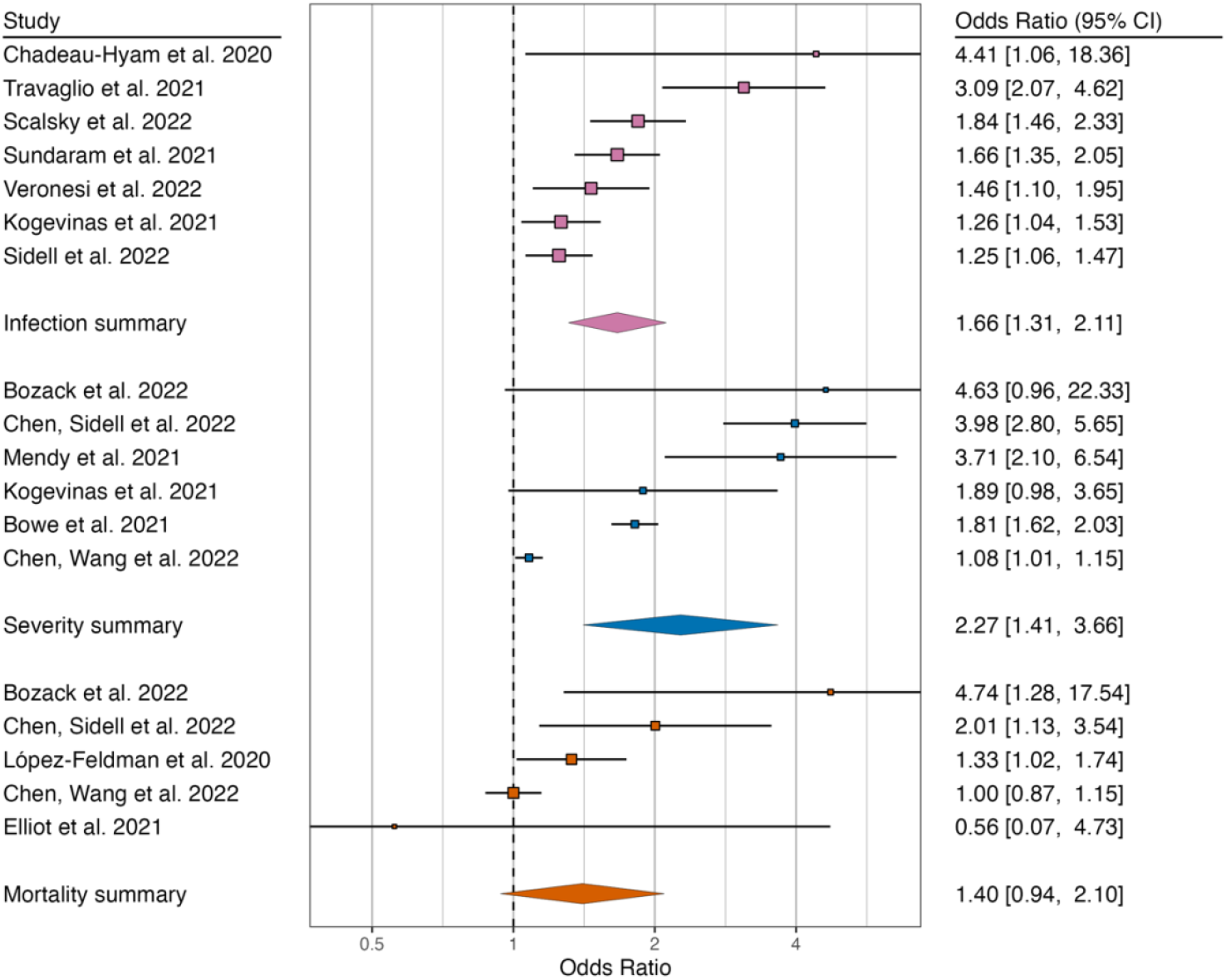
Forest plot of meta-analyses of three outcomes: COVID-19 infection (pink), severity (blue), and mortality (orange) Note: OR represents change in odds of outcome associated with every 10 μg/m^3^ increase in ambient PM_2.5_ exposure. The size of the square represents relative meta-analytic weight of each study.

### Study characteristics

All 18 included studies used a cohort design and focused on background ambient PM_2.5_; none were case-control studies. No study investigated discrete, large-scale PM_2.5_ exposures, meaning we were unable to address our second research question.

Half the studies used North American data (*N* = 9), mostly from the US (*N* = 6), followed by Canada (*N* = 2) and Mexico (*N* = 1). The remainder mostly used European data (*N* = 8), primarily the UK (*N* = 4), followed by Italy (*N* = 2), Spain, and Poland (*N* = 1 each). The last study used Chinese data.

### Study quality

Study quality is summarised in Table 1. Most (13 of 18 studies) were rated “good”. More detail is available in the *Critical appraisal* document on our public repository (9).

**Table 1.**
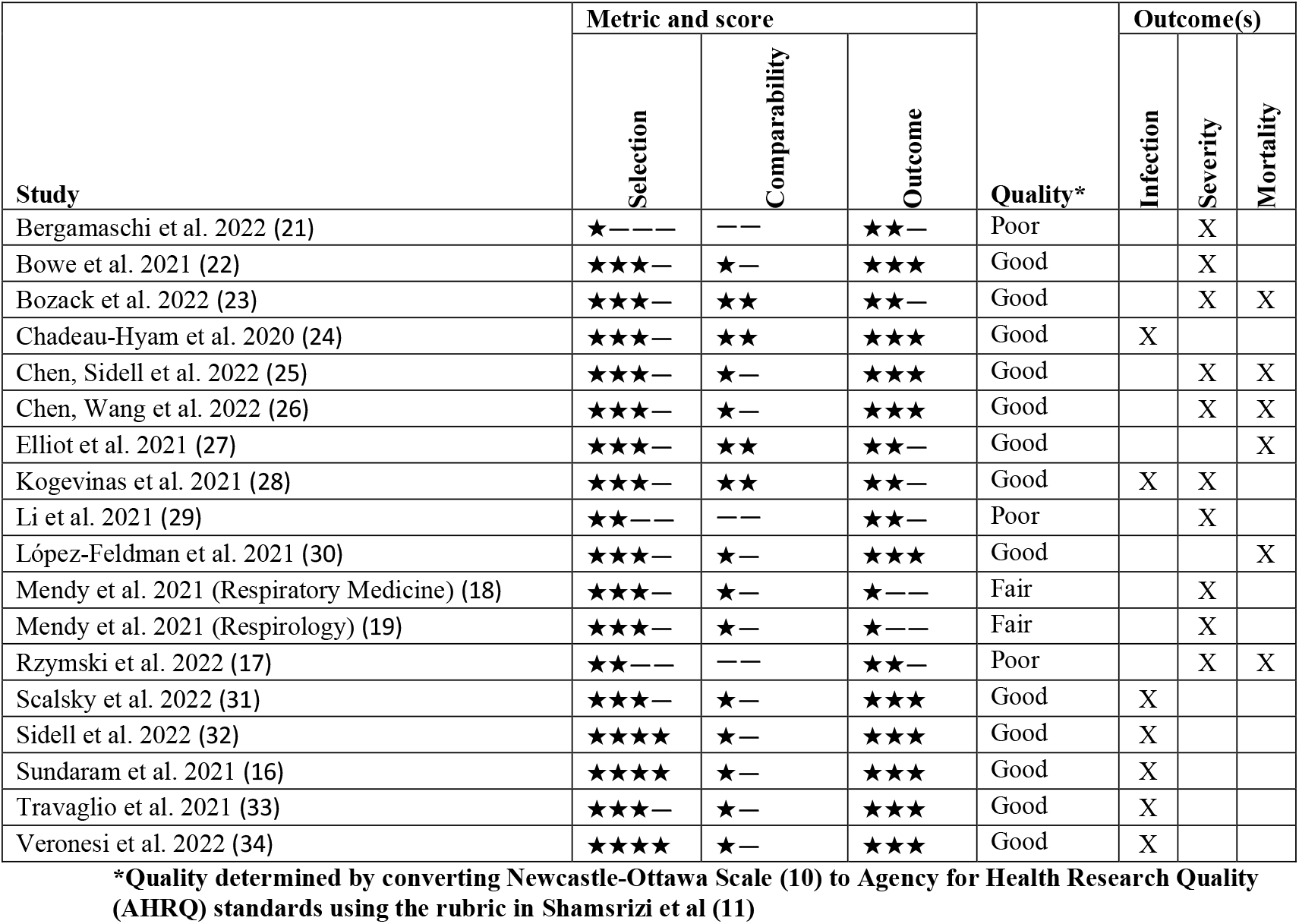
Quality assessment of included studies

Several methodological limitations are worth nothing. Only four of the 18 studies included individual-level adjustments for socioeconomic factors (e.g., education, household income, insurance status). Of the remaining 14, three did not adjust for any socioeconomic factors. Three of the seven infection studies only included participants with a COVID-19 test (24,31,33), while the remainder either used entire cohorts regardless of whether there was a record of a COVID-19 test (32,34) or conducted analyses of the entire cohort as well as just those tested (16,28). Similarly, all but one study examining severity (28) and mortality (27) were limited to cohorts who were diagnosed with COVID-19, while three were restricted to patients hospitalised with COVID-19 (17,23,29). Restricted cohorts present a risk of collider bias, as PM_2.5_ exposure could influence both whether an individual sought testing for COVID-19 or was COVID-19 positive, resulting in distorted associations (35).

Other methodological issues were not captured by the NOS tool. Three studies (16,24,27) included multiple predictors of interest within a single model rather than build models around PM_2.5_ as an exposure. As these are not designed to account for how independent variables may interact (e.g., as mediators or colliders), the statistical associations are less reliable (36).

Some studies reported resolutions up to 100m^2^ (28,31), others used entire cities (29) or monitoring stations spaced tens of kilometres apart (32,37). Several did not specify PM_2.5_ resolution. The timeframe of PM_2.5_ measurement also varied considerably, from just the week prior to inclusion/recruitment (17) up to ten years (18,19) and nearly two decades (30).

All studies using UK data relied on the UK Biobank. While this sample is large at around 500,000 people, it is not considered representative of the UK population due to low participation rates and a skew towards older persons (38,39). Additionally, three used PM_2.5_ estimates from 2010 (24,27,31) – a decade old – and all used participant residences from 2006-2010. This does not account for change of address or the steady decline in PM_2.5_ in the interim (40).

Most studies used single-pollutant models, i.e., PM_2.5_ without any other air pollutants (*N* = 12). The remainder were evenly split between only multi-pollutant models (16,24,27) and both single and multi-pollutant models (29,34,37). Rzymski et al. (17) used a dichotomous indicator of PM_2.5_ based on whether the mean or maximum exceeded 20μg/m^3^ in the week before admission to hospital with COVID-19. All others used continuous mean PM_2.5_, while Mendy et al. (18) also used the maximum.

### PM_2.5_ exposure and COVID-19 infection

Seven studies examined PM_2.5_ and COVID-19 infection, which are summarised in Supplementary Table 1. All were rated “good” quality and reported a significant and positive association. Pooled results indicated a 10μg/m^3^ increase in PM_2.5_ was associated with a 66% increase in the odds of COVID-19 infection (95% CI: 1.31 to 2.11), with 83% of the variance attributable to heterogeneity (*p* < 0.001). Egger’s regression suggested publication bias (*p* = 0.012). Trim-and-fill points could not be applied, though results from leave-one-out sensitivity analysis remained significant with estimates ranging from 1.48 to 1.78 (see Supplementary Figures 1 and 2).

Associations were not consistent across analyses, though no negative association was identified, i.e., all were positive or null. Kogevinas et al. (28) did not find an association with infection determined solely by serological tests within the subsample who agreed to take a test (*n* = 3,922). However, there was a significant association within both the serological test subsample and the full sample (*n* = 9,088) when infection was determined by combining serological tests *and* self-report indicators. The difference may be attributable to limited sensitivity of the serological tests, leading to false-negatives; only 70% of cases identified through self-reported indicators had detectable COVID-19 antibodies.

Sundaram et al. (16) categorised PM_2.5_ exposures into five ordinal categories, which exhibited J-shaped curve with COVID-19 infection; compared to the lowest exposure group (2-6μg/m^3^), COVID-19 infection risk was lower 6-7μg/m^3^, similar (7-8μg/m^3^), and then increasingly higher in the next two groups (8-9μg/m^3^ and ≥10μg/m^3^). This was the case whether the comparisons were between those testing positive for COVID-19 and *not* testing positive (i.e., testers and non-testers in the Ontario population; *N* = 14,695,579) or between those testing positive for COVID-19 and those testing negative (i.e., testers only; *N* = 758,791).

The single-pollutant model that accounted for socioeconomic factors in Veronesi et al. (34) found COVID-19 infections increased 3.6% (95% CI: 1.009-1.075) for every 1μg/m^3^ in PM_2.5_. The single-pollutant model that omitted socioeconomic factors was similar but with a slightly bigger effect (RR: 1.051; 95% CI: 1.027-1.075), which increased substantially when other air pollutants were added to the model (NO_2_: 1.347, NO: 1.105, O_3_: 1.107). This suggests multi-collinearity between air pollutants, which may bias the association between PM_2.5_ and COVID-19 infections. Otherwise, associations remained significantly positive in single and multi-pollutant models.

### PM_2.5_ exposure and COVID-19 severity

Nine studies examined PM_2.5_ and COVID-19 severity, of which five were rated “good”, two “fair”, and two “poor”. These are summarised in Supplementary Table 2. Mendy et al. (18) was the only study not to find a significant association, though in a later study the authors found a significant association when the same participants were included in a substantially larger cohort (*n* = 1,128 versus *n* = 14,783) and PM_2.5_ estimates were updated by a year (19). Aside from Kogevinas et al. (28), all cohorts were restricted to those diagnosed or hospitalised with COVID-19. Severity was indicated in numerous ways including hospitalisation (*N* = 5), ICU admission (*N* = 3), requiring respiratory support (*N* = 3), clinical symptomatology (*N* = 1), oxygen saturation (*N* = 1), or multiple indicators (*N* = 1).

Pooled results from *N* = 6 studies indicate the odds of a severe outcome was 227% higher (95% CI: 1.41 to 3.66) for every 10μm/g^3^ increase in PM_2.5_. Nearly all the variance in effects was due to heterogeneity (*I*^2^: 97%; *p* < 0.001). There was no detectable publication bias (*p* = 0.132). Trim-and-fill points slightly attenuated the results (OR: 2.04; 95% CI: 1.29 to 3.21) and the association remained significant in all leave-one-out analyses (see Supplementary Figure 4).

Chen, Sidell et al. (37) found a consistent associations between PM_2.5_ and COVID-19 hospitalisation when using PM_2.5_ measured over the previous year. The associations were consistent but weaker when using PM_2.5_ from the previous month. This study also found that the association remained when controlling for another air pollutant, NO_2_. Similarly, Li et al. (29) found a positive association between PM_2.5_ and clinically-defined severe COVID-19 across four different lag periods (0-7 days to 0-28 days), which attenuated but remained mostly significant when adjusting for other air pollutants.

### PM_2.5_ exposure and COVID-19 mortality

Five studies examined PM_2.5_ and COVID-19 mortality, which are summarised in Supplementary Table 3. Four of six studies found a significant positive association. The remainder were null. One study was rated “poor”; the rest were “good”.

Pooled results from *n* = 5 studies were positive but non-significant (OR: 1.40; 95% CI: 0.94 to 2.10), with heterogeneity explaining 75% of the variance (*p* = 0.010). There was no evidence of publication bias (*p* = 0.100). Trim-and-fill points could not be applied, though leave-one-out sensitivity analysis indicated the results remained positive but only became significant with the exclusion of Chen, Wang et al (26) (OR: 1.66; 95% CI: 1.06 to 2.59). These results are summarised in Supplementary Figures 5 and 6.

Elliot et al. (27) was the only study that did not restrict its sample to those diagnosed or hospitalised with COVID-19, avoiding associated issues of collider bias. It also had a null finding with a negative point estimate (OR: 0.94, 95% CI: 0.75-1.18). However, the model included multiple predictors rather than being built around a single exposure-outcome relationship, meaning associations were less reliable.

Of the remaining studies, all but Chen, Wang et al. (26) found a significant positive association between PM_2.5_ and COVID-19 mortality. Chen, Sidell et al. (37) found that PM_2.5_ was consistently associated with higher mortality rates across multiple models, regardless of whether PM_2.5_ was measured in the previous month or year and whether the model adjusted for NO_2_.

## Discussion

We found strong evidence that PM_2.5_ exposure increases the risk of COVID-19 infection and weaker evidence that it increases severity and risk of death.

Studies of COVID-19 infection were generally of high quality and consistently demonstrated a significant association with PM_2.5_ across methodologies and populations. Most importantly, the effect was observed even when adjusting for individual-level socioeconomic indicators, probably the most important confounder (4). While there was evidence of publication bias in COVID-19 literature, leave-one-out sensitivity analysis was robust to exclusions.

The association with COVID-19 infection was observed in both single and multi-pollutant models across all studies, suggesting PM_2.5_ is not just an indicator of a generalised effect of poor air quality on risk of COVID-19 infection, but an independent, causal predictor. However, multicollinearity may be an issue, as indicated by increase in effect size when other air pollutants were added to PM_2.5_ models in Veronesi et al. (34).

The evidence on COVID-19 severity and mortality also indicates a positive association, though the quality of the research was weaker and pooled mortality results were non-significant. Nearly every study was limited to people already diagnosed or hospitalised with COVID-19, introducing potential collider bias, or more specifically endogenous selection bias (41). As the above results suggest that PM_2.5_ influences who gets COVID-19, it could also mean that the infected cohorts differ substantially based on their PM_2.5_ exposure. For instance, PM_2.5_ may expand infections into less-vulnerable populations, reducing baseline risk of severe infection and biasing the association with PM_2.5_ towards null.

Kogevinas et al. (28) was the lone COVID-19 severity study to include participants who were not already infected. It also designed statistical models to examine the effect of PM_2.5_ rather than including multiple predictors in a single model, and used high-resolution measures at 100m^2^, finding a positive association with COVID-19 severity. Elliot et al. (27) was the only mortality study to include participants not diagnosed or hospitalised with COVID-19. While it found no association with PM_2.5_ exposure, all predictors were included in a single model, making the results less reliable (36). However, its exclusion in leave-one-out sensitivity analysis did not meaningfully affect pooled results.

Despite the weakness of evidence for effects of PM_2.5_ on COVID-19 severity and mortality, there are still reasons to treat it as real. There is strong circumstantial evidence of a mechanism, including effects of PM_2.5_ on receptor expression, cell defence, and cardiovascular and pulmonary health (3,5,6) which may make infected persons more vulnerable to worse COVID-19 outcomes. Combined with the positive (if not always significant) associations identified in this review, PM_2.5_ air pollution should be treated as a risk factor for severe COVID-19 disease and death.

### Evidence gaps

We identified two major evidence gaps. The first is a lack of cohort or case-control studies of COVID-19 severity and mortality that were not limited to those with COVID-19 and that built models specifically around PM_2.5_ exposure. The second gap is a lack of cohort or case-control studies on discrete, large-scale PM_2.5_ exposures such as smoke from wildfires. It remains unknown whether intensive PM_2.5_ exposure increases short and long-term risks of respiratory illnesses like COVID-19. There is some ecological evidence on an association, though this mainly focuses on concurrent PM_2.5_ exposure (42–44). In the months following the 2019-2020 Black Summer fires in New South Wales, Australia, areas with more burn coverage had higher rates of COVID-19. However, there was no detectable association with larger particulate matter, PM_10_, and the study did not investigate PM_2.5_ (45). We therefore have little idea whether and how long people may be at elevated risk of COVID-19 following major smoke exposures.

### Strengths and limitations

Among this systematic review’s strengths are an inclusion criterion that limited evidence to studies using individual-level data, a quality assessment that indicated most were of good quality, and synthesis of data with a meta-analysis. This review covers studies published in the first 2.5 years of the pandemic, building on previous reviews with more up-to-date evidence.

There are some limitations. Operationalisation of PM_2.5_ exposure varied across studies, including when it was measured, precision, and time periods covered. No studies captured variations in individual exposure due to time spent outdoors or regular movement into areas like the workplace. The review was restricted to outdoor air pollution exposures, which, combined with most studies originating in developed countries, may not be applicable to lower-income countries where indoor air pollution from ‘dirty’ heating and cooking fuels are a greater threat. While all studies used individual-level data, many used aggregated indicators for important confounders like socioeconomic status. COVID-19 is still a relatively new illness, so this review can only be considered an early snapshot of the evidence.

## Conclusion

There is strong evidence that PM_2.5_ increases COVID-19 infections. The evidence for effects on COVID-19 severity and mortality is weaker, but similarly suggests that PM_2.5_ exposure increases risk. When considered alongside evidence that PM_2.5_ worsens cardiovascular and pulmonary health, we see good reason to treat the association with severe illness and death from COVID-19 as real, if not yet fully established. No cohort or case-control studies focused on discrete, large-scale PM_2.5_ exposures such as smoke from wildfires, which will become increasingly important as climate change increases both the frequency and intensity of wildfires.

## Data Availability

All data and materials are available online on a public repository: https://doi.org/10.26180/21529158.v3

## Appendix search terms

The following terms were used to search for relevant studies in Medline, Embase, and the WHO COVID-19 database. In Medline and Embase, terms were searched as MeSH headings. The WHO COVID-19 database does not support MeSH headings, so terms were searched as key words.

> Air pollution OR maximum allowable concentration OR threshold limit values OR petroleum pollution OR traffic-related pollution OR particulate matter OR particulate matter 2.5 OR coal ash OR dust OR smog OR smoke OR soot OR air pollutants OR gasoline OR vehicle emissions OR particle pollution
>
> AND
>
> COVID-19 OR SARS-CoV-2 OR coronavirus disease 2019 OR severe acute respiratory syndrome coronavirus 2
>
> AND
>
> Virulence OR patient acuity OR severity of illness index OR morbidity OR basic reproduction number OR incidence OR prevalence OR mortality OR fatal outcome OR survival rate OR death OR hospitalisation OR length of stay OR patient admission OR asymptomatic diseases OR asymptomatic infections OR critical illness

---

**Supplementary Table 1.**
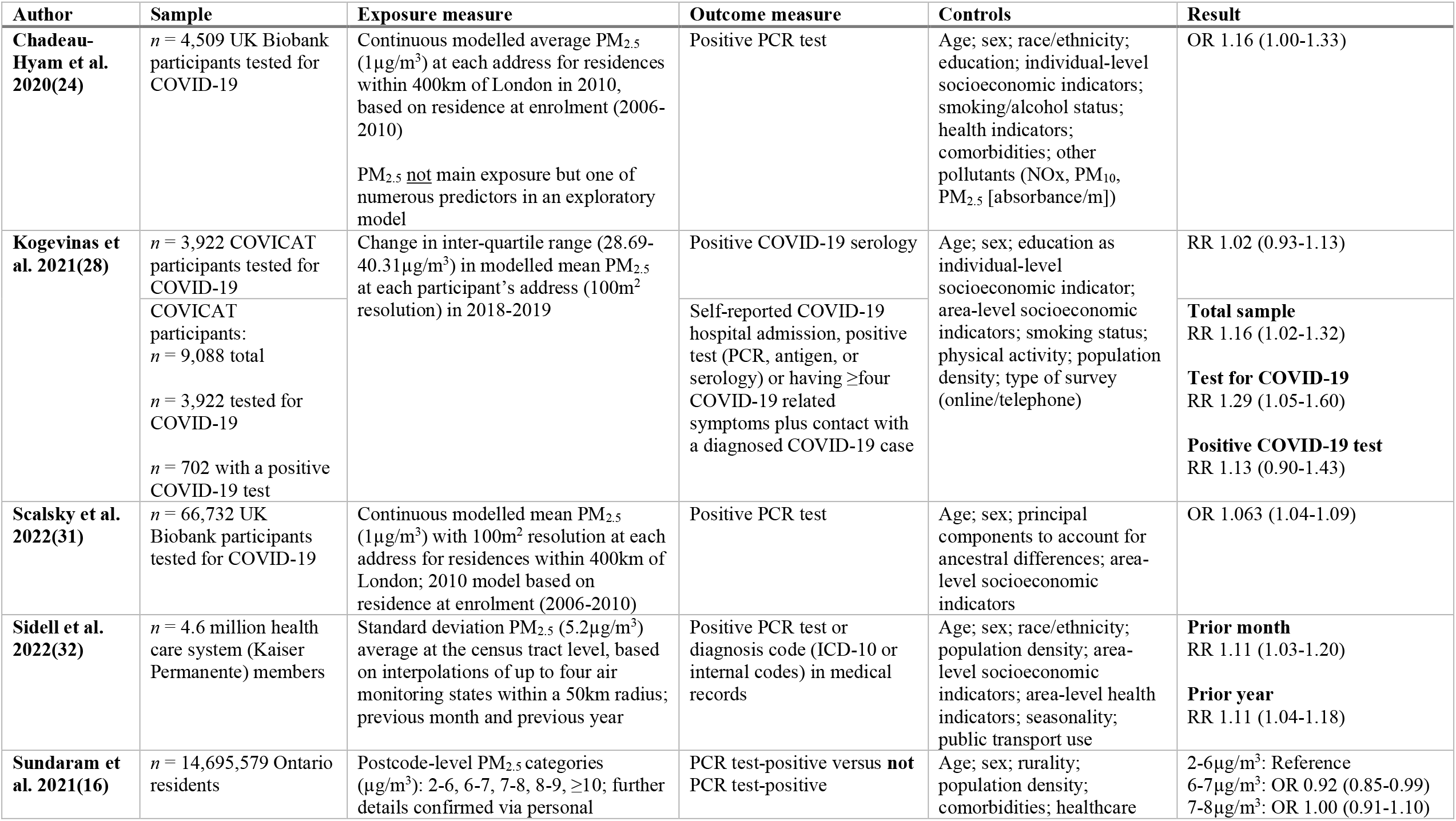
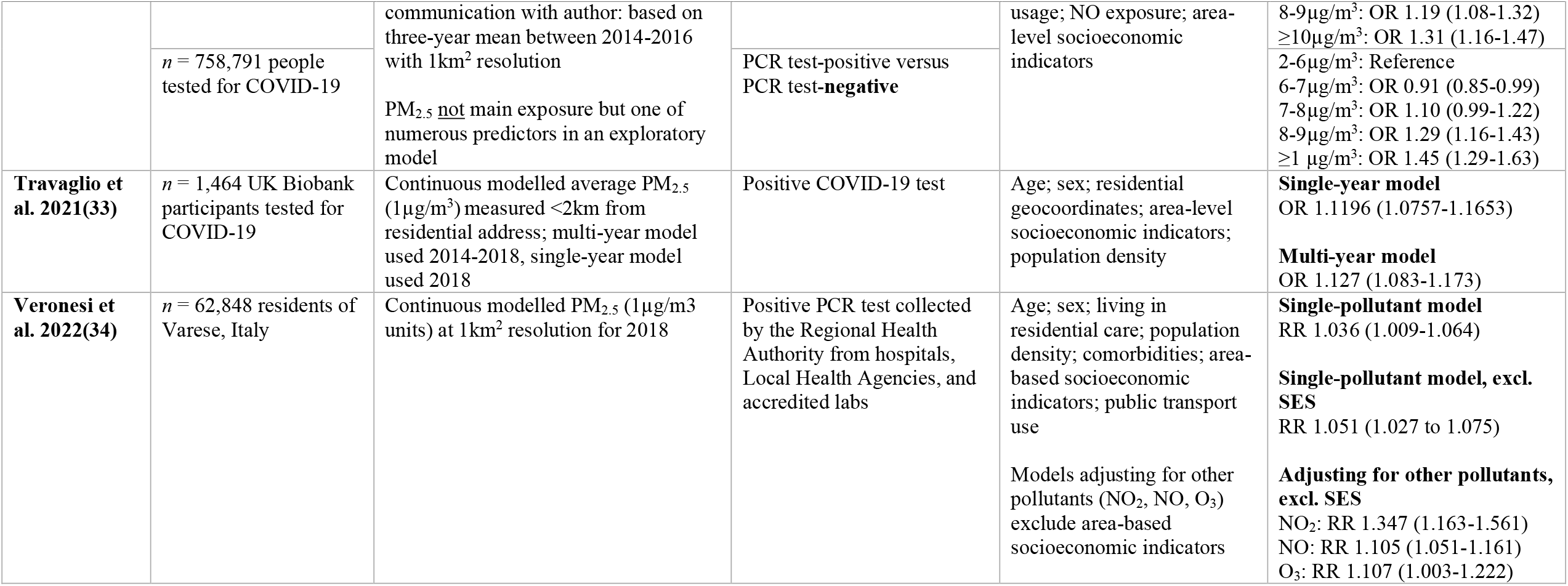
Characteristics of included studies which examine the relationship between PM_2.5_ exposure and COVID-19 infection.

**Supplementary Table 2.**
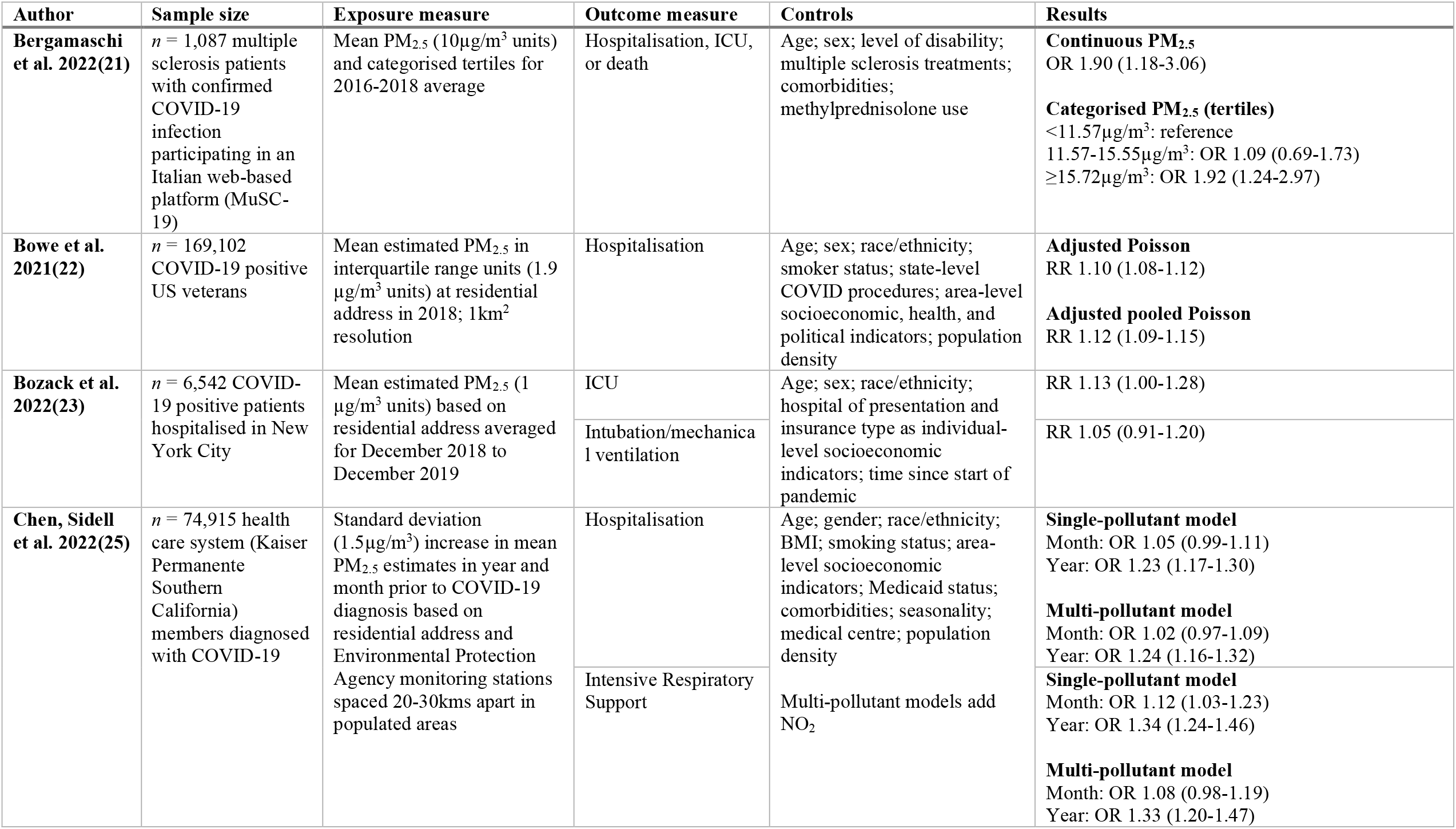
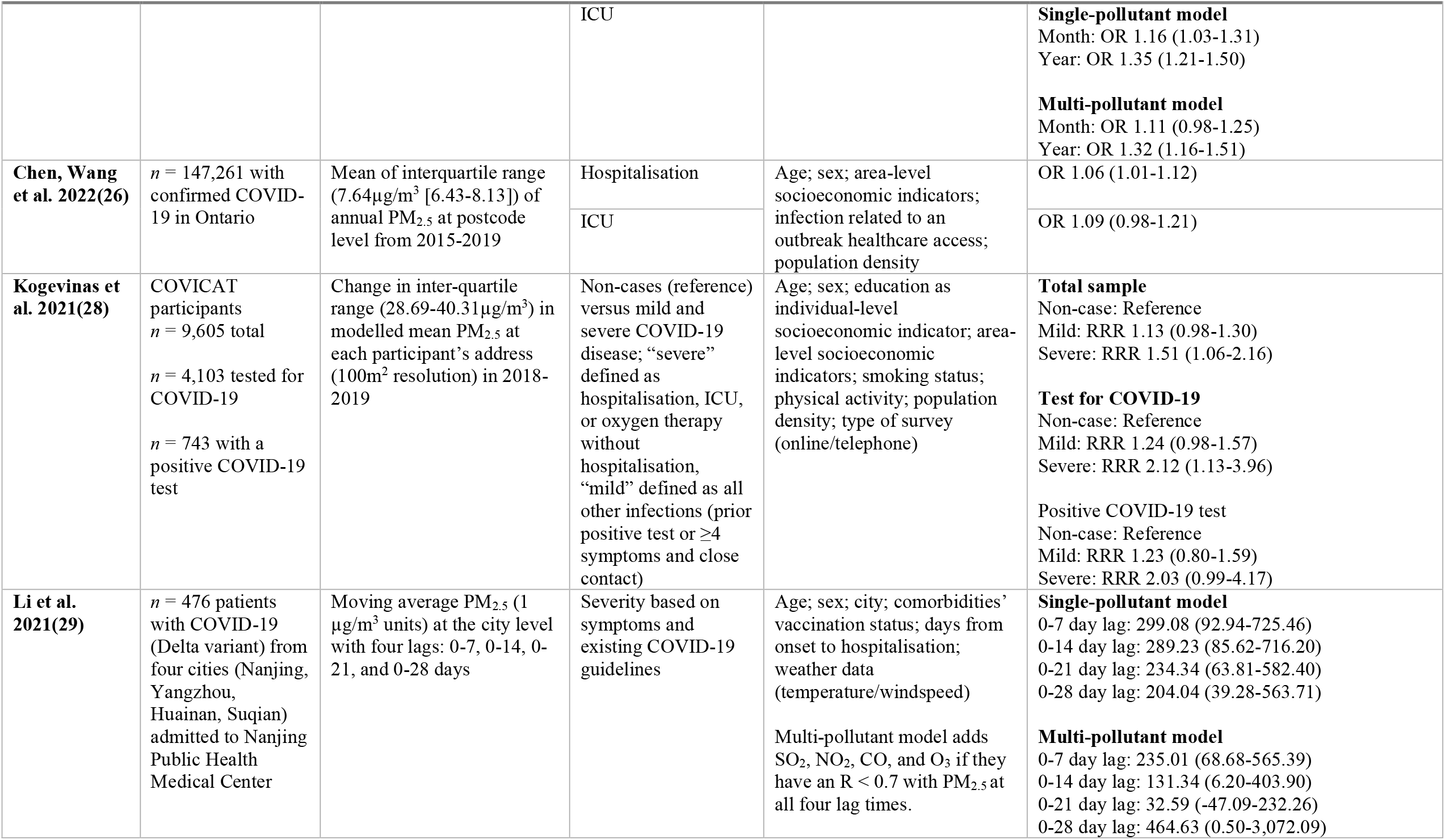
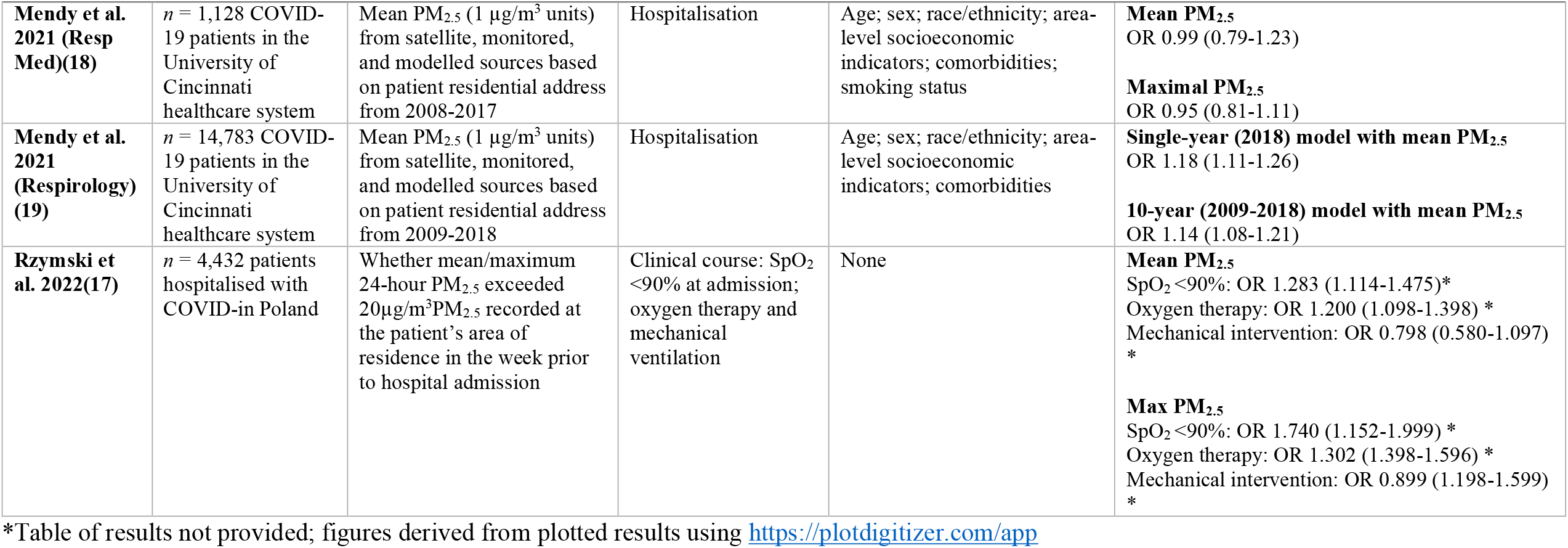
Characteristics of included studies which examine the relationship between PM_2.5_ exposure and COVID-19 severity.

**Supplementary Table 3.**
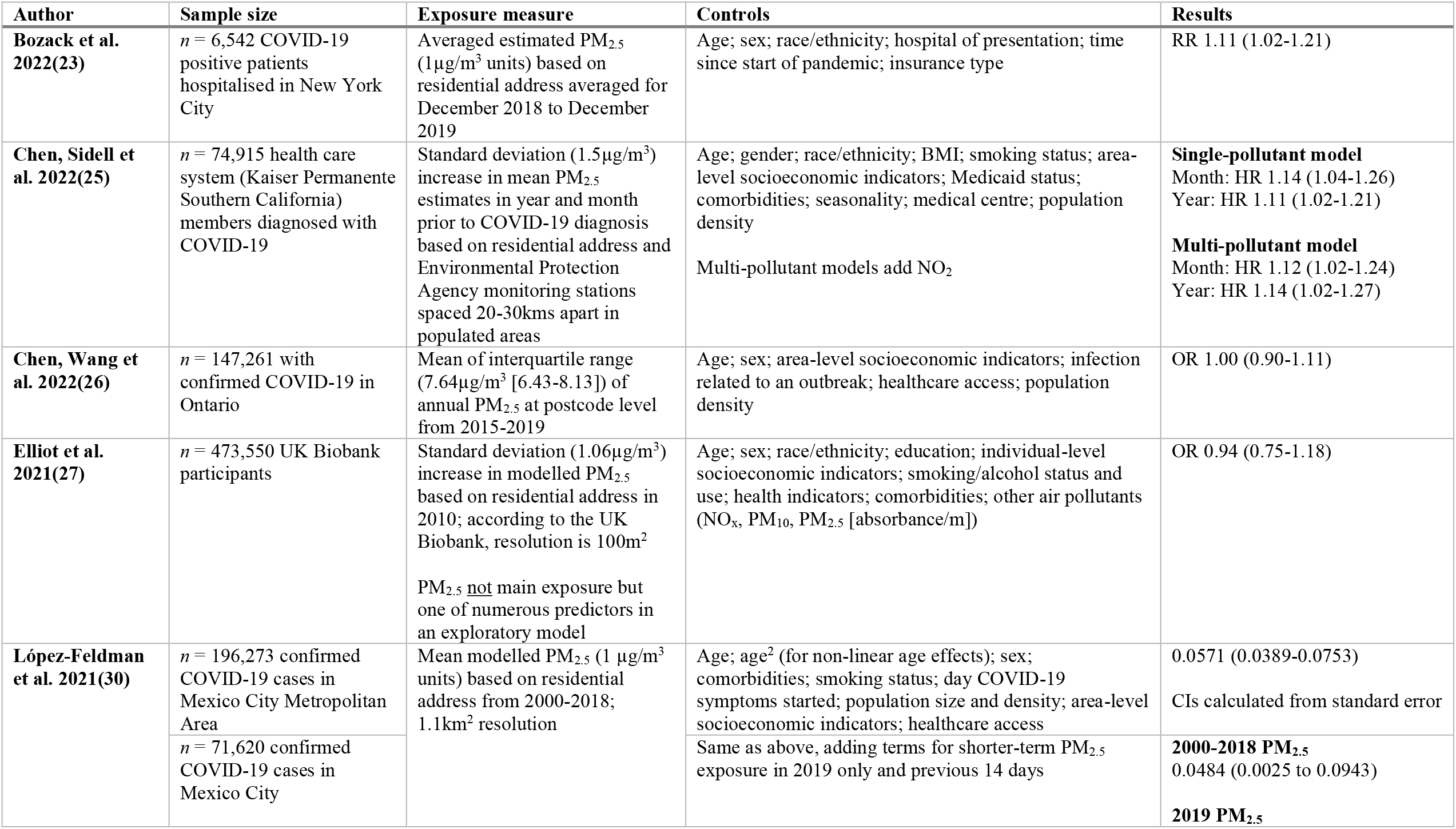
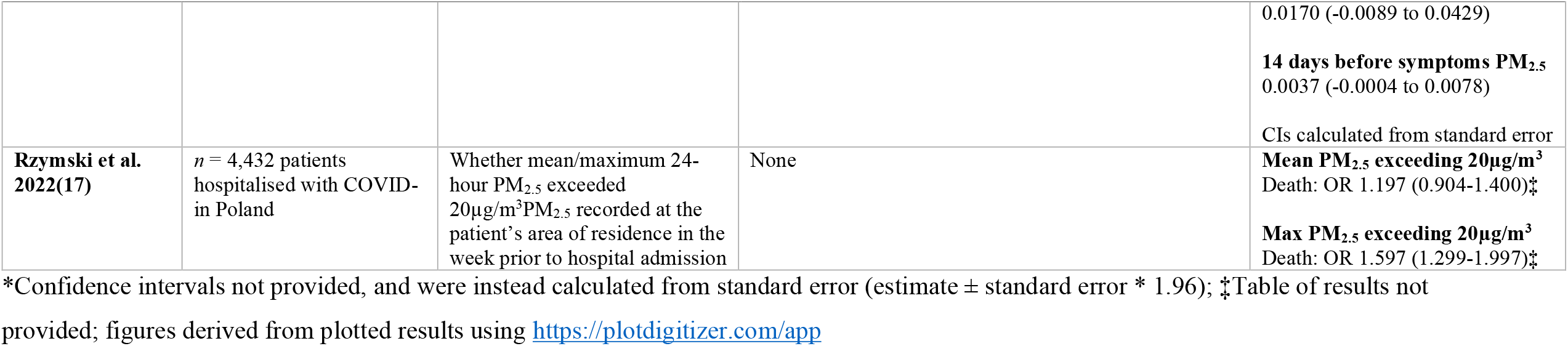
Characteristics of included studies which examine the relationship between PM_2.5_ exposure and COVID-19 mortality.

**Supplementary Figure 1.**
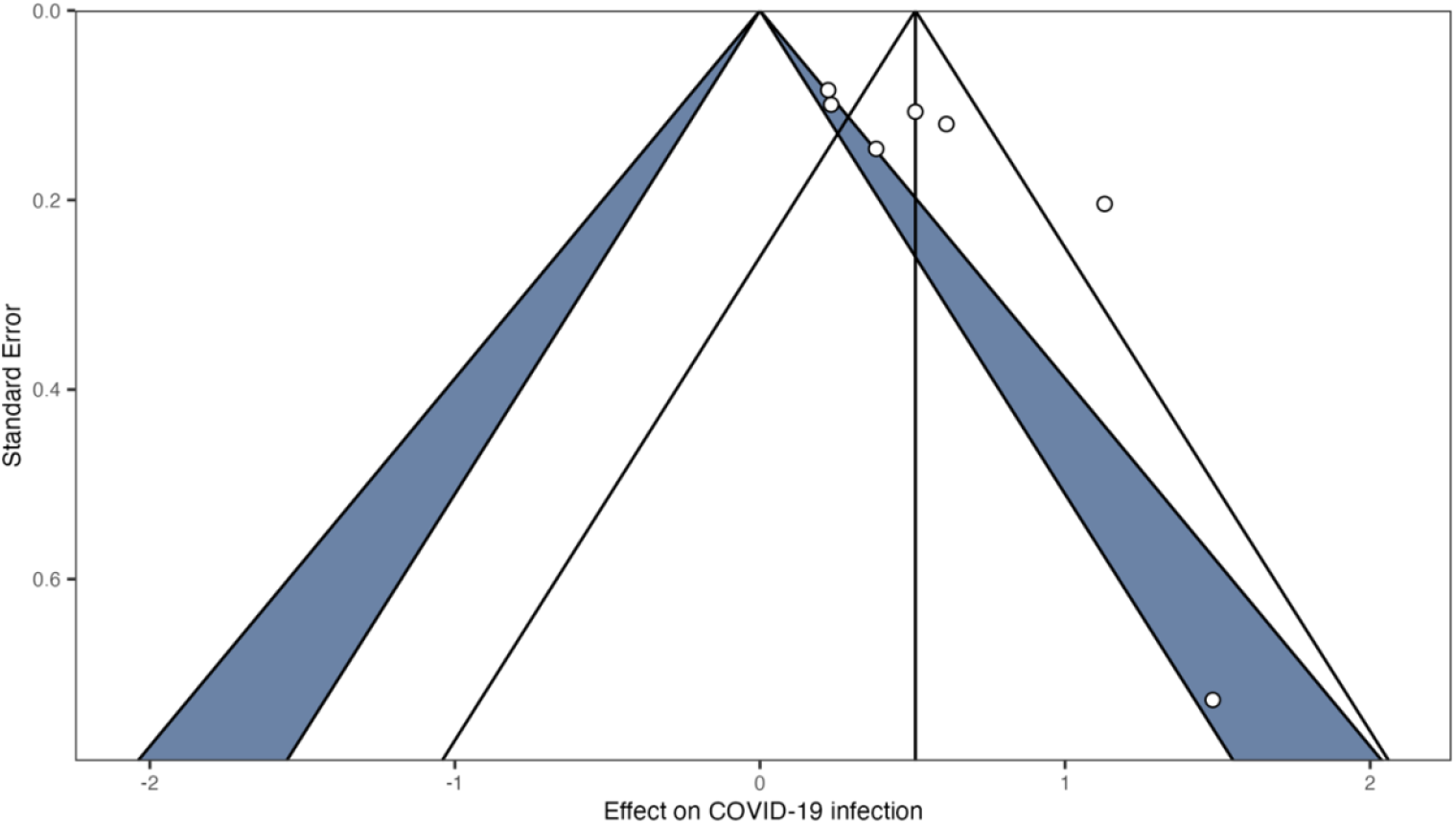
Funnel plot for PM_2.5_ and COVID-19 infection studies.

**Supplementary Figure 2.**
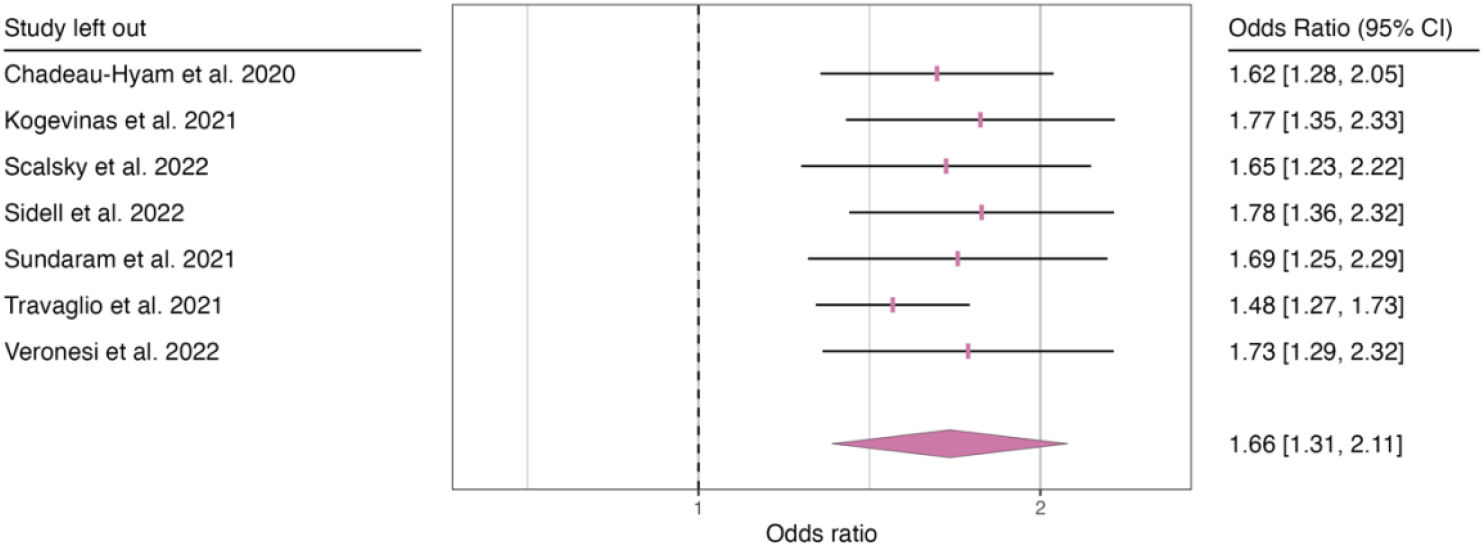
Leave-one-out sensitivity analysis for PM_2.5_ and COVID-19 infection. Note: OR represents change in odds of outcome associated with every 10 μg/m^3^ increase in ambient PM_2.5_ exposure. The size of the square represents relative meta-analytic weight of each study.

**Supplementary Figure 3.**
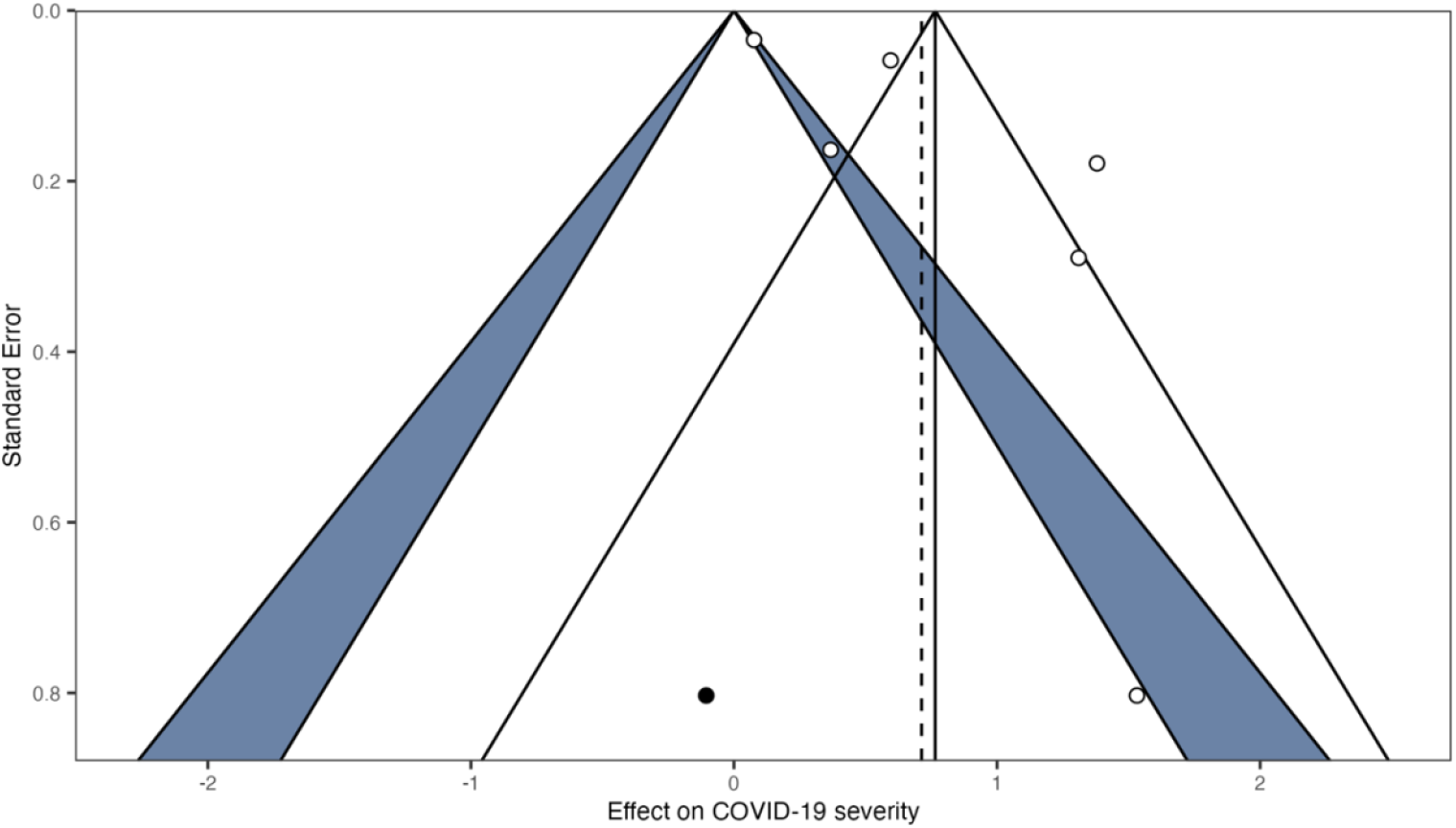
Funnel plot for PM_2.5_ and COVID-19 severity studies (black points are trim- and-fill points to account for publication bias)

**Supplementary Figure 4.**
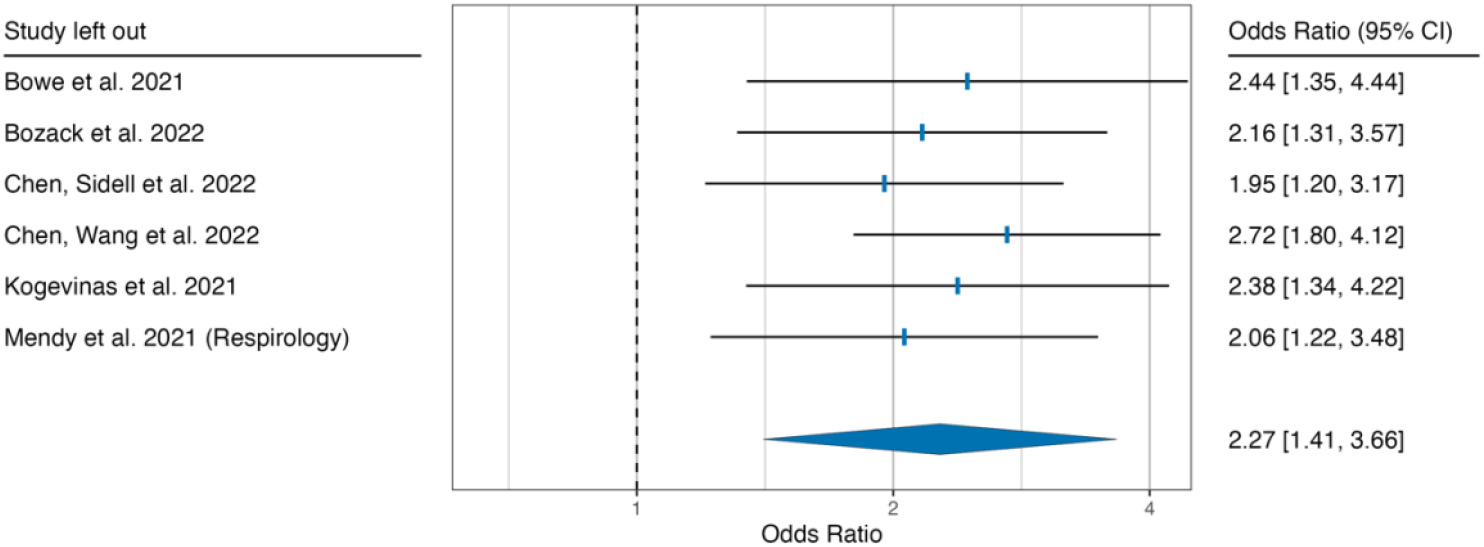
Leave-one-out sensitivity analysis for PM_2.5_ and COVID-19 severity. Note: OR represents change in odds of outcome associated with every 10 μg/m^3^ increase in ambient PM_2.5_ exposure. The size of the square represents relative meta-analytic weight of each study.

**Supplementary Figure 5.**
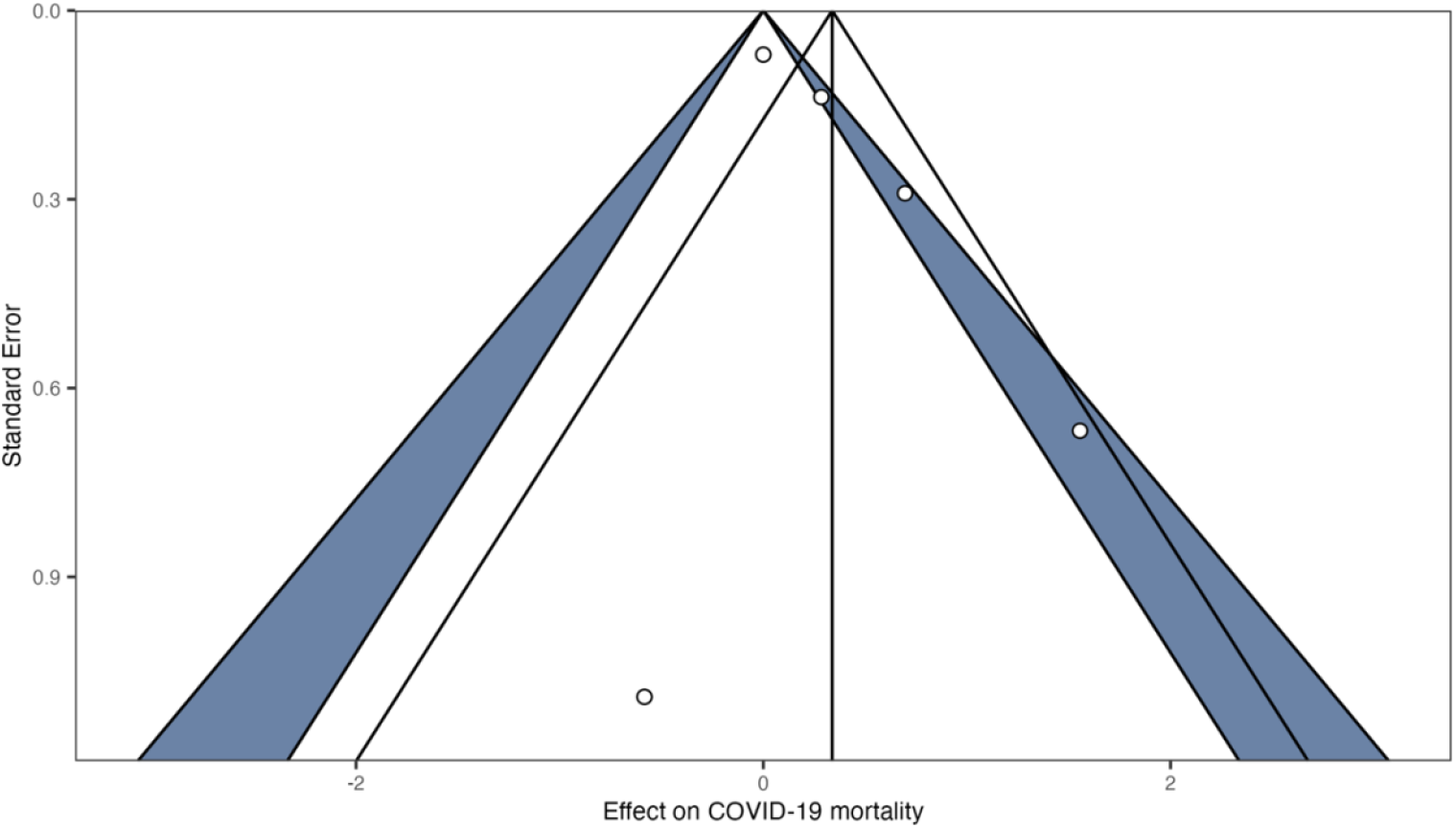
Funnel plot for PM_2.5_ and COVID-19 mortality studies.

**Supplementary Figure 6.**
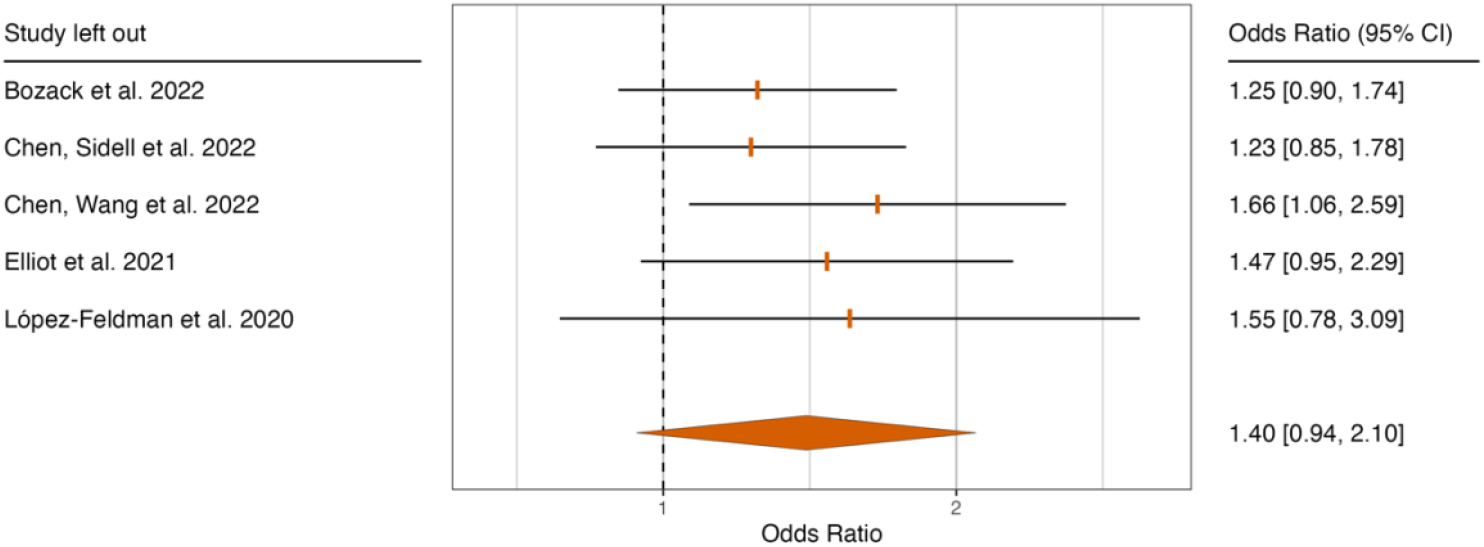
Leave-one-out sensitivity analysis for PM_2.5_ and COVID-19 severity. Note: OR represents change in odds of outcome associated with every 10 μg/m^3^ increase in ambient PM_2.5_ exposure. The size of the square represents relative meta-analytic weight of each study.

**Supplementary Figure 7.**
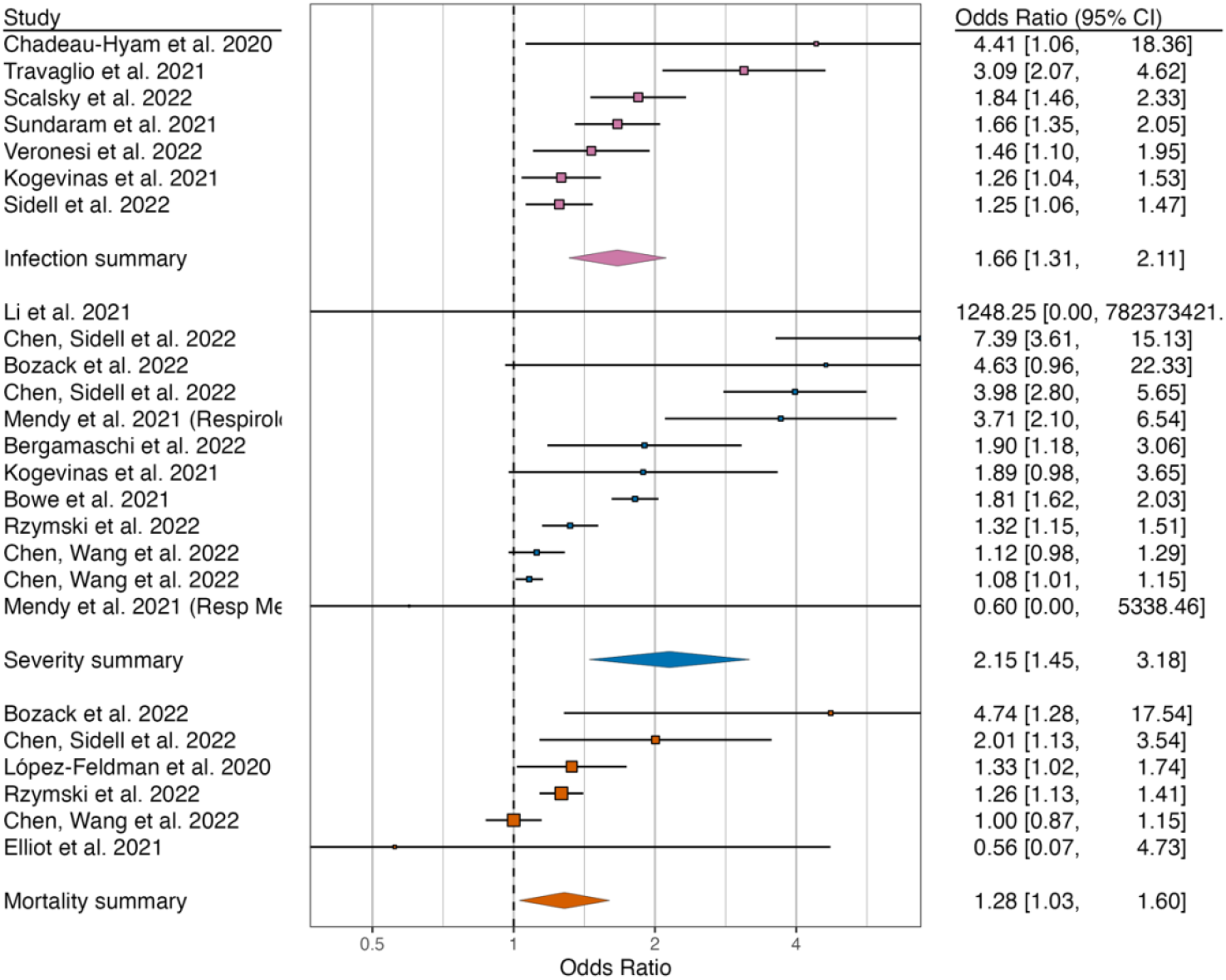
Meta-analysis change in odds of COVID-19 infection (pink), severity (blue), and mortality (orange) for every 10μg/m3 of mean PM_2.5_regardless of quality rating. Note: OR represents change in odds of outcome associated with every 10 μg/m^3^ increase in ambient PM_2.5_ exposure. The size of the square represents relative meta-analytic weight of each study.

## Notes

### Competing Interest Statement

The authors have declared no competing interest.

### Clinical Protocols

https://www.crd.york.ac.uk/prospero/display_record.php?ID=CRD42022345129

### Funding Statement

This study did not receive any funding.

### Author Declarations

This is a systematic review and meta-analysis, using only published research results. These may not be widely available to the public due to paywalls.

## References

1. Katoto PDMC, Brand AS, Bakan B, Obadia PM, Kuhangana C, Kayembe-Kitenge T, et al. Acute and chronic exposure to air pollution in relation with incidence, prevalence, severity and mortality of COVID-19: a rapid systematic review. Environ Health. 2021 Dec;20(1):41.

2. Copat C, Cristaldi A, Fiore M, Grasso A, Zuccarello P, Signorelli SS, et al. The role of air pollution (PM and NO_2_) in COVID-19 spread and lethality: A systematic review. Environmental Research. 2020 Dec;191:110129.

3. Bourdrel T, Annesi-Maesano I, Alahmad B, Maesano CN, Bind MA. The impact of outdoor air pollution on COVID-19: a review of evidence from in vitro, animal, and human studies. Eur Respir Rev. 2021 Mar 31;30(159):200242.

4. Blakely T, Hunt D, Woodward A. Confounding by socioeconomic position remains after adjusting for neighbourhood deprivation: an example using smoking and mortality. Journal of Epidemiology & Community Health. 2004 Dec 1;58(12):1030–1.

5. Rebuli ME, Brocke SA, Jaspers I. Impact of inhaled pollutants on response to viral infection in controlled exposures. Journal of Allergy and Clinical Immunology. 2021 Dec;148(6):1420–9.

6. Landrigan PJ, Fuller R, Acosta NJR, Adeyi O, Arnold R, Basu N (Nil), et al. The Lancet Commission on pollution and health. The Lancet. 2018 Feb;391(10119):462–512.

7. Sheppard N, Lane TJ, Carroll M. A systematic review examining the relationship between PM2.5 exposure and risk of COVID-19-related morbidity and mortality at an individual level [Internet]. PROSPERO; 2022. Available from: https://www.crd.york.ac.uk/prospero/display_record.php?ID=CRD42022345129

8. Page MJ, McKenzie JE, Bossuyt PM, Boutron I, Hoffmann TC, Mulrow CD, et al. The PRISMA 2020 statement: an updated guideline for reporting systematic reviews. BMJ. 2021 Mar 29;372:n71.

9. Gao CX, Lane TJ, Sheppard N, Carroll M. PM2.5 and COVID-19 systematic review [Internet]. Bridges. 2022. Available from: https://doi.org/10.26180/21529158.v2

10. Wells GA, Shea B, O’Connell D, Peterson J, Welch V, Losos M, et al. The Newcastle-Ottawa Scale (NOS) for assessing the quality of nonrandomised studies in meta-analyses [Internet]. 2013. Available from: https://www.ohri.ca/programs/clinical_epidemiology/oxford.asp

11. Shamsrizi P, Gladstone BP, Carrara E, Luise D, Cona A, Bovo C, et al. Variation of effect estimates in the analysis of mortality and length of hospital stay in patients with infections caused by bacteria-producing extended-spectrum beta-lactamases: a systematic review and meta-analysis. BMJ Open. 2020 Jan;10(1):e030266.

12. Viechtbauer W. Conducting meta-analyses in R with the metafor package. Journal of Statistical Software. 2010;36(3):1–48.

13. Kossmeier M, Tran US, Voracek M. metaviz: Forest Plots, Funnel Plots, and Visual Funnel Plot Inference for Meta-Analysis [Internet]. 2019. Available from: https://cran.r-project.org/package=metaviz

14. R Core Team. R: A language and environment for statistical computing [Internet]. Vienna, Austria: R Foundation for Statistical Computing; 2022. Available from: https://www.r-project.org/

15. Stroup DF, Berlin JA, Olkin I, Williamson GD, Rennie D, Moher D, et al. Meta-analysis of Observational Studies in Epidemiology: A proposal for reporting. JAMA. 2000 Apr 19;283(15):2008–12.

16. Sundaram ME, Calzavara A, Mishra S, Kustra R, Chan AK, Hamilton MA, et al. Individual and social determinants of SARS-CoV-2 testing and positivity in Ontario, Canada: a population-wide study. CMAJ. 2021 May 17;193(20):E723–34.

17. Rzymski P, Poniedzialek B, Rosinska J, Rogalska M, Zarębska-Michaluk D, Rorat M, et al. The association of airborne particulate matter and benzo[a]pyrene with the clinical course of COVID-19 in patients hospitalized in Poland. Environmental Pollution. 2022 Aug;306:119469.

18. Mendy A, Wu X, Keller JL, Fassler CS, Apewokin S, Mersha TB, et al. Long-term exposure to fine particulate matter and hospitalization in COVID-19 patients. Respiratory Medicine. 2021 Mar;178:106313.

19. Mendy A, Wu X, Keller JL, Fassler CS, Apewokin S, Mersha TB, et al. Air pollution and the pandemic: Long-term PM_2.5_ exposure and disease severity in COVID-19 patients. Respirology. 2021 Dec;26(12):1181–7.

20. Rzymski P, Poniedzialek B, Rosinska J, Ciechanowski P, Peregrym M, Pokorska-Spiewak M, et al. Air pollution might affect the clinical course of COVID-19 in pediatric patients. Ecotoxicology and Environmental Safety. 2022 Jul;239:113651.

21. Bergamaschi R, Ponzano M, Schiavetti I, Carmisciano L, Cordioli C, Filippi M, et al. The effect of air pollution on COVID-19 severity in a sample of patients with multiple sclerosis. Euro J of Neurology. 2022 Feb;29(2):535–42.

22. Bowe B, Xie Y, Gibson AK, Cai M, van Donkelaar A, Martin RV, et al. Ambient fine particulate matter air pollution and the risk of hospitalization among COVID-19 positive individuals: Cohort study. Environment International. 2021 Sep;154:106564.

23. Bozack A, Pierre S, DeFelice N, Colicino E, Jack D, Chillrud SN, et al. Long-term air pollution exposure and COVID-19 mortality: A patient-level analysis from New York City. Am J Respir Crit Care Med. 2022 Mar 15;205(6):651–62.

24. Chadeau-Hyam M, Bodinier B, Elliott J, Whitaker MD, Tzoulaki I, Vermeulen R, et al. Risk factors for positive and negative COVID-19 tests: a cautious and in-depth analysis of UK biobank data. International Journal of Epidemiology. 2020 Oct 1;49(5):1454–67.

25. Chen Z, Sidell MA, Huang BZ, Chow T, Eckel SP, Martinez MP, et al. Ambient air pollutant exposures and COVID-19 severity and mortality in a cohort of COVID-19 patients in Southern California. Am J Respir Crit Care Med. 2022 May 10;rccm.202108-1909OC.

26. Chen C, Wang J, Kwong J, Kim J, van Donkelaar A, Martin RV, et al. Association between long-term exposure to ambient air pollution and COVID-19 severity: a prospective cohort study. CMAJ. 2022 May 24;194(20):E693–700.

27. Elliott J, Bodinier B, Whitaker M, Delpierre C, Vermeulen R, Tzoulaki I, et al. COVID-19 mortality in the UK Biobank cohort: revisiting and evaluating risk factors. Eur J Epidemiol. 2021 Mar;36(3):299–309.

28. Kogevinas M, Castaño-Vinyals G, Karachaliou M, Espinosa A, de Cid R, Garcia-Aymerich J, et al. Ambient air pollution in relation to SARS-CoV-2 infection, antibody response, and COVID-19 disease: A cohort study in Catalonia, Spain (COVICAT Study). Environ Health Perspect. 2021 Nov 17;129(11):117003.

29. Li Z, Tao B, Hu Z, Yi Y, Wang J. Effects of short-term ambient particulate matter exposure on the risk of severe COVID-19. Journal of Infection. 2022 May;84(5):684–91.

30. López-Feldman A, Heres D, Marquez-Padilla F. Air pollution exposure and COVID-19: A look at mortality in Mexico City using individual-level data. Science of The Total Environment. 2021 Feb;756:143929.

31. Scalsky RJ, Chen YJ, Ying Z, Perry JA, Hong CC. The social and natural environment’s impact on SARS-CoV-2 infections in the UK Biobank. IJERPH. 2022 Jan 4;19(1):533.

32. Sidell MA, Chen Z, Huang BZ, Chow T, Eckel SP, Martinez MP, et al. Ambient air pollution and COVID-19 incidence during four 2020–2021 case surges. Environmental Research. 2022 May;208:112758.

33. Travaglio M, Yu Y, Popovic R, Selley L, Leal NS, Martins LM. Links between air pollution and COVID-19 in England. Environmental Pollution. 2021 Jan;268:115859.

34. Veronesi G, De Matteis S, Calori G, Pepe N, Ferrario MM. Long-term exposure to air pollution and COVID-19 incidence: a prospective study of residents in the city of Varese, Northern Italy. Occup Environ Med. 2022 Mar;79(3):192–9.

35. Griffith GJ, Morris TT, Tudball MJ, Herbert A, Mancano G, Pike L, et al. Collider bias undermines our understanding of COVID-19 disease risk and severity. Nat Commun. 2020 Dec;11(1):5749.

36. Westreich D, Greenland S. The Table 2 Fallacy: Presenting and interpreting confounder and modifier coefficients. American Journal of Epidemiology. 2013 Feb 15;177(4):292–8.

37. Chen Z, Sidell MA, Huang BZ, Chow T, Eckel SP, Martinez MP, et al. Ambient air pollutant exposures and COVID-19 severity and mortality in a cohort of patients with COVID-19 in southern California. Am J Respir Crit Care Med. 2022 Aug 15;206(4):440–8.

38. Keyes KM, Westreich D. UK Biobank, big data, and the consequences of non-representativeness. The Lancet. 2019 Mar;393(10178):1297.

39. Swanson JM. The UK Biobank and selection bias. The Lancet. 2012 Jul;380(9837):110.

40. Department for Environment Food & Rural Affairs. Concentrations of particulate matter (PM10 and PM2.5) [Internet]. GOV.UK. 2022 [cited 2022 Nov 16]. Available from: https://www.gov.uk/government/statistics/air-quality-statistics/concentrations-of-particulate-matter-pm10-and-pm25

41. Elwert F, Winship C. Endogenous selection bias: The problem of conditioning on a collider variable. Annual Review of Sociology. 2014;40:31–53.

42. Mace Firebaugh C, Beeson T, Wojtyna A, Arboleda R. Increase PM_2.5_ levels associated with increased incidence of COVID-19: The Washington wildfires of 2020. Environ Smoke. 2021 Aug 31;4(2):49–53.

43. Meo SA, Abukhalaf AA, Alomar AA, Alessa OM, Sami W, Klonoff DC. Effect of environmental pollutants PM-2.5, carbon monoxide, and ozone on the incidence and mortality of SARS-COV-2 infection in ten wildfire affected counties in California. Science of The Total Environment. 2021 Feb;757:143948.

44. Zhou X, Josey K, Kamareddine L, Caine MC, Liu T, Mickley LJ, et al. Excess of COVID-19 cases and deaths due to fine particulate matter exposure during the 2020 wildfires in the United States. Sci Adv. 2021 Aug 13;7(33):eabi8789.

45. Cortes-Ramirez J, Michael RN, Knibbs LD, Bambrick H, Haswell MR, Wraith D. The association of wildfire air pollution with COVID-19 incidence in New South Wales, Australia. Science of The Total Environment. 2022 Feb;809:151158.

